# Transdiagnostic gray matter volume reductions implicating social dysfunction in depression, schizophrenia, and alcohol use disorder: A neuroimaging meta-analysis

**DOI:** 10.64898/2026.10.18.26365862

**Authors:** Mercy Chepngetich Bore, Jingxian He, Mengfan Han, Abraham Tonny Hagan, Xiqin Liu, Jie Zhang, Keith Kendrick, Xiaoqi Huang, Juan Yang, Benjamin Klugah-Brown, Benjamin Becker

**Author notes:** **Corresponding authors:** Benjamin Becker, Email address; Postal/mail address: University of Hong Kong, Pokfulam, Hong Kong, China / University of Electronic Science and Technology of China, Xiyuan Ave 2006, 611731 Chengdu, China; Benjamin Klugah-Brown, Email address; Postal/mail address: University of Electronic Science and Technology of China, Xiyuan Ave 2006, 611731 Chengdu, China.

## Abstract

Anhedonia, social withdrawal and executive deficits represent transdiagnostic symptoms in depression, schizophrenia and addiction, yet transdiagnostic brain volumetric alterations and their contribution to these symptom domains have not been systematically characterized. Here, we capitalized on a pre-registered (https://osf.io/s57uq) coordinate-based meta-analysis across 147 VBM studies (n=13,041) to determine common and disorder-specific gray matter deficits in patients with schizophrenia (90 studies), major depression disorder (44 studies) and alcohol use disorder (13 studies). Original case-control articles comparing patients with healthy controls, published in English and reporting whole-brain results were included. We found shared GMV alterations in the ventromedial prefrontal cortex, ventral anterior cingulate cortex, insula, superior temporal gyrus. On the network level, we found robust functional interactions of the identified shared regions with the cortical midline structures as well as social and theory of mind-related behavioral functions. Subsequent characterization analyses highlighted the involvement of dopaminergic and serotonergic neurotransmitters as well as associated genetic annotations. Overall, the present meta-analysis found common as well as separate GMV abnormalities in schizophrenia, major depressive disorder and alcohol use disorder suggesting a common neurostructural substrate and treatment target for social withdrawal and anhedonia across these disorders.

## Introduction

Anhedonia – the diminished capacity to experience pleasure or interest in previously rewarding activities – represents a key transdiagnostic symptom across major depressive disorder (MDD), schizophrenia (SZ), and alcohol use disorder (AUD) [1, 2]. Together with transdiagnostic deficits in executive [3, 4] and social functioning [4, 5], these impairments contribute substantially to the chronic course, long-term functional disability and poor treatment response in these conditions. These transdiagnostic symptoms are not responsive to the established interventions [6, 7] and determining the underlying neural basis could help to determine targeted novel interventions for the transdiagnostic dysfunctions.

Schizophrenia is a chronic mental disorder associated with negative symptoms including anhedonia and asociality. With a lifetime prevalence of approximately 1%, it continues to contribute to the global burden of disease [8]. The first positive symptoms typically emerge after the development of cognitive and negative symptoms [9]. Previous neuroimaging meta-analyses have suggested volumetric decrease in patients with schizophrenia in several networks encompassing the temporolimbic structures, insula and striatal regions [10]. Major depressive disorder is often characterized by depressed mood and anhedonia (prolonged loss of interest in almost all daily activities). Some neurobiological models have been proposed to account for MDD pathogenesis including the role of dysfunctional cortico-limbic networks in MDD [11, 12]. Previous VBM meta-analyses have found gray matter decreases in patients with MDD in regions encompassing the anterior cingulate cortex, frontal gyrus, hippocampus and insula [13–15].

Alcohol use disorder (AUD) is highly prevalent substance use disorder that carries a huge global health burden and causes more than 2 million deaths each year [16, 17]. Individuals with AUD often suffer from compulsive use of alcohol despite the negative social and cognitive consequences that arise [18]. Numerous studies have documented gray matter reductions among individuals with AUD compared to healthy controls. Furthermore, previous VBM meta-analyses on AUD suggested significantly reduced gray matter in extensive regions of the prefrontal cortex, striatal and insular regions, anterior cingulate cortex (ACC) as well as parts of the posterior cingulate cortex (PCC) [19–21].

While MDD, SZ and AUD present unique symptoms and clinical phenotypes, accumulating evidence indicates shared neurobiological substrates. The transdiagnostic symptom deficits in anhedonia, cognition and social domains face substantial treatment challenges [22, 23]. Anhedonia in these disorders has been closely linked to dysregulation of mesolimbic reward circuitry, particularly dopaminergic projections from the midbrain to the ventral striatum and ventromedial prefrontal cortex (vmPFC) [24, 25]. Converging evidence from disorder-specific studies indicates dysregulations in dopaminergic functioning and reward-related functioning of the striatum and vmPFC in MDD, SZ and AUD [26–29].

Executive functioning deficits including working memory and inhibition deficits have been identified as major endophenotype candidates across the disorders and patients with these disorders have consistently demonstrated dysfunctional prefrontal cortex engagement during executive tasks [30–33]. Social dysfunctions including empathy and theory of mind (ToM) have been hypothesized to represent a trans-diagnostic symptomatology domain [34, 35] alterations in these domains have been linked to functional and structural alterations in the vmPFC in depression, schizophrenia and AUD [5, 35–37]. However, the neural basis of the transdiagnostic deficits and their associations with the behavioral domains have not been systematically determined.

Neuroimaging meta-analyses have been increasingly adopted as the gold standard for solving heterogeneity and generalizability problems in neuroimaging research [38, 39] as well as determine transdiagnostic brain structural signatures [13, 40, 41]. The current meta-analysis aimed to determine common and separate gray matter volume alterations in schizophrenia, depression and alcohol use disorder patients compared to healthy controls to determine transdiagnostic alterations that underly anhedonia, executive deficits and social dysfunctions. Specifically, we aimed to (a) Determine disorder-specific GMV deficits between patients and healthy controls within disorders, (b) Determine the common gray matter deficits in schizophrenia, major depression disorder and alcohol use disorder (c) Employ advanced network, behavioral, and molecular-genetic analyses to dissect the contribution of these alterations to the three transdiagnostic symptom clusters. In line with previous literature, we hypothesized that all three disorders would exhibit decreased brain volumes in prefrontal regions involved in executive control and value processing. We further hypothesized that schizophrenia patients would show gray matter decreases in the thalamus, amygdala and hippocampus, depression patients would exhibit GMV decreases in prefrontal brain regions while alcohol use disorder would show GMV alterations in frontostriatal brain regions.

## Methods

The present meta-analysis followed standard procedures of coordinate-based meta-analyses and the Preferred Reporting Items for Systematic Reviews and Meta-Analyses (PRISMA) guidelines [42, 43]. To enhance credibility and interpretability of the results, the meta-analysis was pre-registered on the Open Science Framework repository prior to data analysis (https://osf.io/s57uq). Of note, we initially did not pre-register the inclusion of AUD group given that the idea only developed when screening the literature for MDD and SZ.

### Literature Search and Study Selection

A comprehensive literature search using three databases of life sciences; PubMed (https://www.ncbi.nlm.nih.gov/pubmed), Web of Science (https://www.webofscience.com/wos/alldb/basic-search) and, PsycInfo https://www.apa.org/pubs/databases/psycinfo/). The search terms were: (schizophrenia OR schizophrenic disorders; depression; alcohol OR alcohol use disorder AND (voxel-based morphometry, OR VBM OR gray matter OR voxelwise) AND (Morphometry OR Voxel-based OR voxelwise).

### Selection Criteria

Two independent reviewers (M.C.B and J.H) screened the eligible literature according to the pre-registered inclusion criteria and any disagreements during the selection process was resolved by a consensus by B.B. Original VBM studies on SZ, MDD and AUD were screened, and suitable studies were identified according to the inclusion and exclusion criteria. Additionally, suitable studies from other sources that fulfill the inclusion criteria were included. The inclusion criteria were: (a) Original peer-reviewed studies, written in English and published between January 2000 and March 2024, (b) studies reporting whole-brain results in standard stereotactic space (Talairach or Montreal Neurological Institute), (c) case-control studies comparing patients and healthy controls, (d) adult participants between 18 to 60 years old. The exclusion criteria will be as follows: (a) studies focusing entirely on participants <18 or those >60 years old, (b) studies focusing on other mental/mood disorders, (c) studies presenting ROI results only, (d) studies including findings from the same dataset as already included studies. The exclusion criteria included: (a) studies reporting region-of-interest results (ROI) only, (b) articles involving participants with comorbid conditions, (c) studies including findings from the same dataset as already included studies.

### Data Extraction

Important demographic, clinical and statistical information was extracted from eligible studies. The data extraction process including authors’ names, titles of articles, year of publication, mean age, number of participants, medication information, etc., was conducted extensively prior to data analysis.

### Statistical Analysis

The meta-analysis was performed using SDM-PSI toolbox version 6.2.1 (https://www.sdmproject.com). The steps are summarized as follows: (a) Peak coordinates of between-group differences (patients versus healthy subjects) and their corresponding effect sizes (t/z values) were extracted. Z-values were converted to t-values using the standard statistical converter (https://www.sdmproject.com/utilities/?show=Coordinates). (b) Preprocessing was performed to estimate the lower and upper bounds (Hedges’ g) of the most probable effect size images [38]. (c) The mean was calculated using maximum-likelihood estimation (MLE) and meta-analysis of non-statistically significant unreported effects (MetaNSUE) algorithms. This step estimates the most likely effect sizes generating several imputations according to estimates within the bounds [44]. (d) Next, imputed study images were recreated. (e) Finally, the permutation test evaluated the combined meta-analysis images for statistical significance [38].

Results of the meta-analysis were visualized by the MRIcroGL (NITRC: MRIcroGL: Tool/Resource Info). The analyses were considered significant at p < .0025 uncorrected, with a voxel-level threshold of k=10 voxels. This threshold balances Type I and Type II errors and has been employed in previous meta-analyses [13, 27, 45].

### Characterization on the network, behavioral, genetic and receptor levels

In line with previous meta-analyses on depression and transdiagnostic disorder alterations, we employed standard procedures according to our previous studies [26, 35]. Network level analyses were achieved by performing seed-based functional connectivity analysis on Data Processing Assistant for Resting-State fMRI (DPARSF) on the MATLAB platform. It was performed to show the important cognitive networks of the core regions of decreased GMV across disorders. Here, peak coordinates of the common regions were used as nodes of separable networks. Data from 100 healthy subjects from the OASIS dataset were included and the corresponding preprocessing steps was run on Data Processing Assistant for Resting-State fMRI (http://rfmri.org/DPARSF) with the following steps: (a) Removal of the first 5 volumes of the time-series, (b) Motion correction using rigid body rotation and translation, while excluding subjects with a maximum motion > 2 mm or 2°, (c) Normalization into standard stereotactic space, which included co-registration to and the matching segmentation matrix was applied to the functional time series. Peak coordinates of the identified clusters were used to determine whether these clusters are part of common or distinct functional large-scale networks. Sphere-based 6mm radius centered seed regions at the identified shared coordinates were used for the voxel-wise functional connectivity analyses. A radius of 6mm was selected because it provides statistically robust network analyses according to previous studies [46]. Next, behavioral level characterization was employed using the Neurosynth database (https://www.neurosynth.org/). This analysis highlighted the specific behavioral functions associated with the common regions. Thirdly, genetic underpinnings of identified regions were identified using the Brain annotation (BAT) toolbox. For this analysis, we extracted the top ten genes in order of their regional expression values and plotted the results. These genetic annotations enabled the identification of risk factors associated with psychiatric disorders.

Finally, we explored the roles of monoamine neurotransmitters i.e., dopamine transporter (DAT) and serotonin (5HT_1b_) which possess both functional and clinical importance in the pathogenesis of common psychiatric disorders as well as a significant role in motivation and reward processing disruptions in these disorders. Brain structure has been associated with neurotransmitter transporters/receptors. DAT was selected because it plays a vital role in adjusting dopamine availability and subsequent dopamine-mediated behaviors while 5HT_1b_ was selected on the basis of its neurological roles of mood, aggression, memory, stress sensibility and anxiety. Receptor-level analysis was therefore conducted to compare the mean densities of dopamine and serotonin receptors in the common regions of gray matter deficits in the three disorders. Receptor characterization followed these steps: First, Positron Emission Tomography (PET) atlases of dopamine and serotonin were acquired (https://xtra.nru.dk/FS5ht-atlas/, https://www.nitrc.org/projects/spmtemplates). Next, reference regions (occipital lobe and cerebellum) were removed by WFU Pickatlas. 6mm sphere-shaped ROIs were created on MarsBaR region of interest toolbox (http://marsbar.sourceforge.net). The corresponding mean values of the two neurotransmitters in the common brain regions of the three disorders were extracted using the Cognitive and Affective Neuroscience Lab (CANlab) Core toolbox (https://canlabweb.colorado.edu/).

### Heterogeneity, sensitivity and publication bias

Tests for heterogeneity were performed on the SDM-PSI platform and examined by the *I^2^* index. Jackknife sensitivity analysis was conducted on Anisotropic Effect Size-Signed Differential Mapping (AES-SDM) to measure the robustness of the meta-analysis findings. Publication bias was evaluated statistically and interpreted by Egger’s tests and funnel plots.

### Exploratory analyses – meta-regression and linear model analysis

We conducted meta-regression analyses to evaluate the potential confounding effects of gender (number of females) and age across the three disorders. A linear model analysis was performed to assess medication effects in SZ and MDD group, but not in AUD due to limited data. Here, medication and non-medication studies were coded as 1 and 0 respectively. A further sub-group meta-analysis of non-medication studies in the two disorders was performed to validate the original meta-analytic findings.

## Results

### Demographic and clinical information of included studies

We included a total of 147 VBM studies encompassing 44 MDD [47–91], 90 Schizophrenia [37, 92–180] and 13 AUD [181–193] studies. A total of n=13,041 participants comprising of 7731 SZ patients and healthy controls, 4618 MDD patients and healthy controls and 692 AUD patients and healthy controls were included in the present meta-analysis. The analysis involved patient data of SZ (n=3812, mean age=32.18, *SD*=8.19), MDD (n=2361, mean age=37.39, *SD*=9.98) and AUD (n=331, mean age=45.15, *SD*=7.83) and healthy controls data from the SZ group (n=3919, mean age=32.08, *SD=*7.94), MDD group (n=2257, mean age=36.66, *SD*=10.21) and AUD group (n=361, mean age=43.7, *SD*=8.17). The flow diagram of the selection process and workflow of the main analyses are displayed in **Fig. 1**. Demographic and clinical characteristics are included in **Supplementary Tables 2, 3, 4.**

**Fig. 1.**
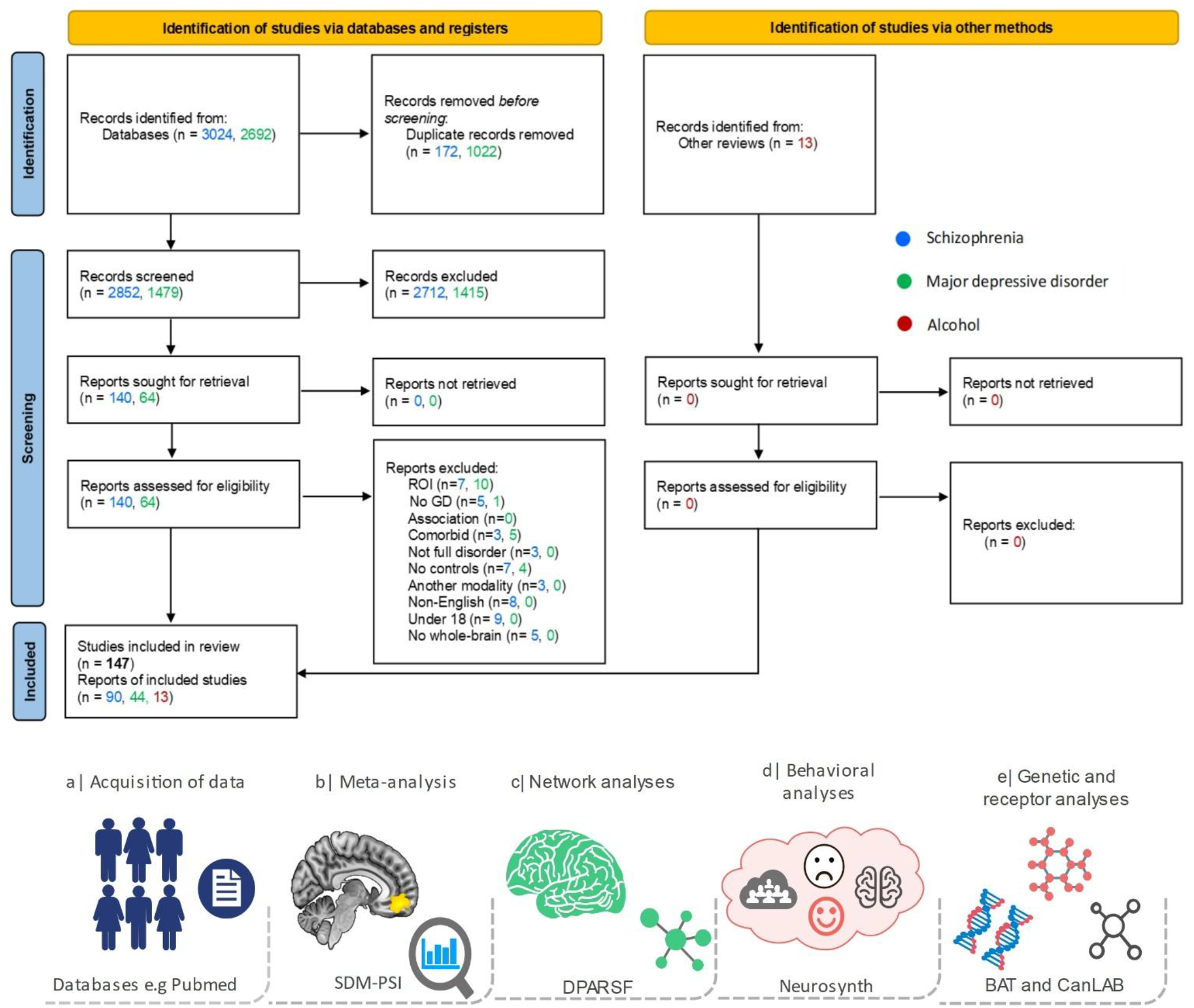
PRISMA flow diagram of the selection process. The workflow of the main analyses conducted is shown below the Prisma flowchart.

### Results of the VBM meta-analysis of SZ, MDD and AUD

The present meta-analysis revealed robustly decreased GMV across meta-analyses. We initially determined alterations within each disorder and found that compared to the healthy reference groups: a) MDD patients demonstrated reduced GMV in the ventromedial prefrontal cortex (vmPFC), mid temporal gyrus and left insula, b) Patients with SZ showed decreased GMV in widely distributed network encompassing the bilateral insula, vmPFC and thalamus. c) Individuals with AUD exhibited reduced GMV largely in the vmPFC, bilateral insula and cerebellum. There were no GMV increases in any of the three disorders (**Fig. 2a-c**). Next, we determined shared alterations by a quantitative comparison analysis of the meta-analytic findings and found four regions exhibiting commonly decreased gray matter across the disorders, i.e., vmPFC extending to the ventral anterior cingulate cortex (vACC), insula and superior temporal gyrus (STG) (**Fig. 2d**). Main meta-analytic findings are displayed in **Table 1** and **Supplementary Table 5** for the disorder-specific clusters.

**Fig. 2.**
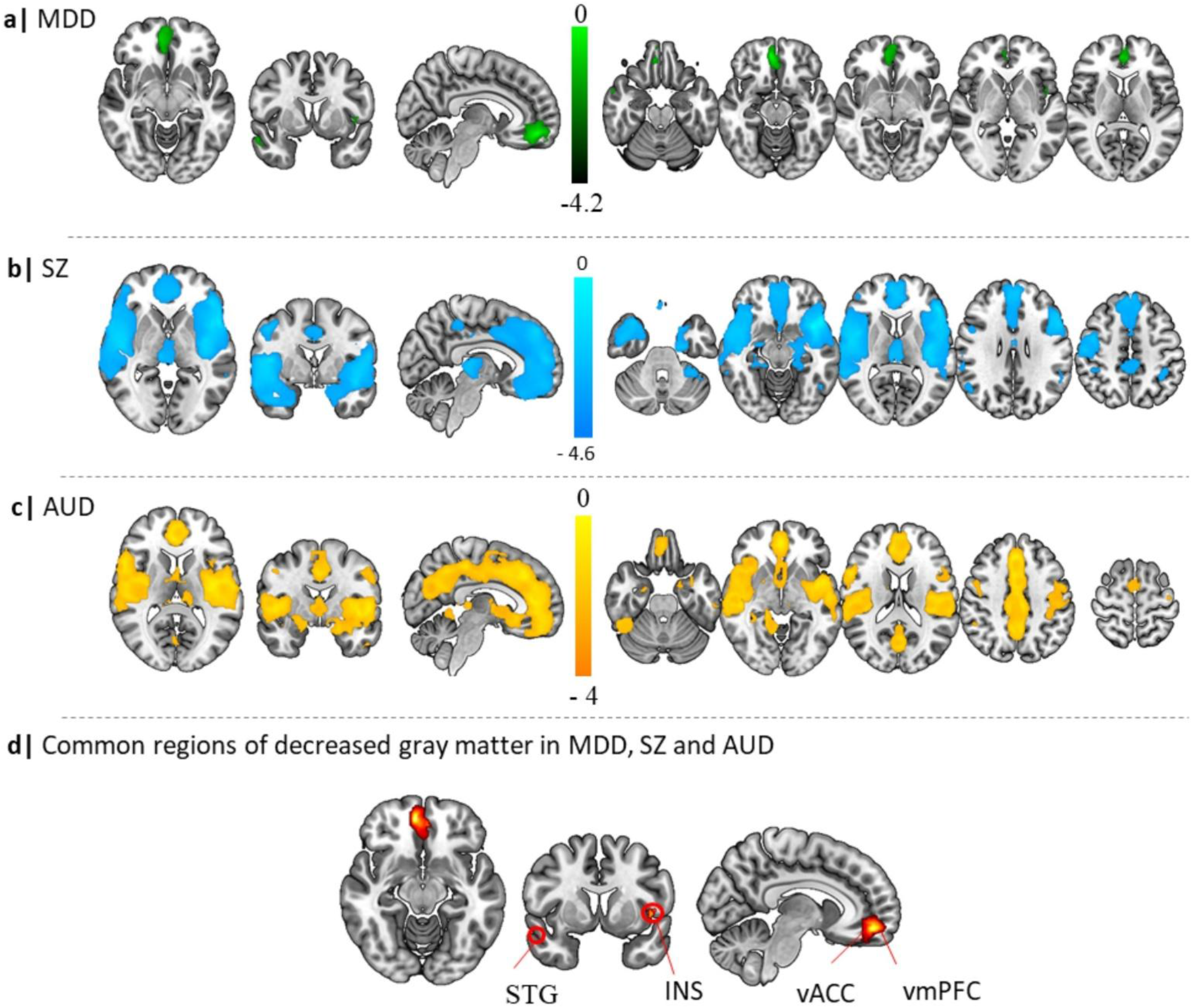
Results of the main meta-analytic findings of Schizophrenia, Major depressive disorder and Alcohol Use disorder compared to healthy controls. **a** Decreased GMV in MDD versus healthy controls, **b** GMV decreases in Schizophrenia patients compared with healthy controls, **c** displays gray matter decreases in individuals with Alcohol Use Disorder versus healthy controls. **d** shows the regions of common gray matter decreases across the three disorders (*p* < .0025, k ≥ 10 voxels).

**Table 1.** Whole-brain meta-analytic results for SZ, MDD and AUD at uncorrected p≤0.0025.

| Contrast | Main brain regions | MNI coordinates | SDM Z | I<sup>2</sup> statistic (%) | Egger's bias | Egger's p | Jackknife sensitivity |
|---|---|---|---|---|---|---|---|
| MDD < HC | R vmPFC | 6, 50, -12 | -5.079 | 1.9 | -0.48 | 0.46 | 44/44 |
|  | R mid temporal gyr | 52, 2, -26 | -3.812 | 18.81 | -0.43 | 0.56 |  |
|  | L insula/ROL | -46, 8, 0 | -3.395 | 42.31 | -0.89 | 0.3 |  |
| MDD > HC |  |  | *None* |  |  |  |  |
| SZ < HC | R insula | 46, 0, 10 | -8.953 | 38.69 | -0.55 | 0.35 | 90/90 |
|  | L insula | -40, 10, -12 | -11.273 | 18.36 | -0.5 | 0.32 |  |
|  | L vmPFC | 2, 46, 40 | -8.071 | 21.83 | -0.57 | 0.25 |  |
|  | L thalamus | -4, -18, 10 | -6.142 | 36.62 | -0.85 | 0.14 |  |
|  | L median cingulate | -6, -36, 50 | -4.311 | 49.62 | -0.41 | 0.51 |  |
|  | R angular gyr | 52, -58, 34 | -4.112 | 25.51 | -0.28 | 0.56 |  |
|  | L IPG | -38, -46, 48 | -4.476 | 44.25 | -0.36 | 0.57 |  |
|  | L cereb | -6, -84, -16 | -4.097 | 17.48 | -0.47 | 0.37 |  |
|  | R inf temporal gyr | 50, -52, -20 | -3.727 | 17.51 | -0.6 | 0.23 |  |
|  | R med cingulate | 10, -12, 42 | -3.620 | 57.15 | -0.78 | 0.23 |  |
|  | L mid temporal gyr | -64, -18, -20 | -3.264 | 0.96 | -0.26 | 0.57 |  |
|  | Corpus callosum | -2, -10, 28 | -3.285 | 62.4 | -0.38 | 0.62 |  |
| SZ > HC |  |  | *None* |  |  |  |  |
| AUD < HC | R vmPFC | 4, 32, 38 | -7.622 | 9.62 | -1.42 | 0.47 | 13/13 |
|  | L insula/ROL | -44, -10, 6 | -7.853 | 5.29 | -1.4 | 0.46 |  |
|  | R insula/ROL | 50, 4, 4 | -7.195 | 1.99 | -1.24 | 0.5 |  |
|  | L Cereb | -34, -52, -48 | -4.710 | 4.34 | -1.13 | 0.57 |  |
|  | R ILF | 40, -52, -14 | -4.87 | 9.2 | -2.19 | 0.29 |  |
|  | Unknown | 16, 0, -14 | -4.335 | 5.44 | 0.91 | 0.67 |  |
|  | R Precentral Gyr | 46, -14, 52 | -4.445 | 9.52 | -1.43 | 0.52 |  |
|  | R Cereb | 20, -68, -50 | -3.753 | 6.53 | -1.07 | 0.63 |  |
|  | R PHG | 32, -36, -10 | -4.448 | 8.47 | -2.29 | 0.27 |  |
|  | L sup parietal gyr | -20, -64, 54 | -3.747 | 7.77 | 0.41 | 0.84 |  |
|  | L inf temporal gyr | -52, -2, -38 | -3.451 | 5.3 | -0.81 | 0.72 |  |
|  | L mid frontal gyr | -46, 30, 32 | -3.115 | 12.54 | -1.12 | 0.66 |  |
|  | BA 48 | 38, 0, -16 | -3.651 | 0.26 | -0.73 | 0.72 |  |
| AUD>HC |  |  | *None* |  |  |  |  |

Abbreviations: AUD, Alcohol Use Disorder, Cereb, cerebellum, Gyr, gyrus, HC, healthy controls, ILF, inferior longitudinal fasciculus, inf, inferior, L, left, MDD, major depressive disorder, MNI, Montreal Neurological Institute, PHG, parahippocampal gyrus, R, right, ROL, Rolandic operculum, SDM, seed-based d mapping, sup, superior, SZ, Schizophrenia, TFCE, threshold-free cluster enhancement, VBM, voxel-based morphometry, vmPFC, ventromedial prefrontal cortex.

### Characterization of VBM alterations on the network, behavioral, genetic and receptor level

We characterized the identified common regions on the network, behavioral, genetic and receptor levels. Network analyses demonstrate the voxel-wise functional connectivity patterns of the common core regions of GMV decreases. Network patterns of the vmPFC revealed strong functional interactions of this region with cortical midline regions such as the frontal pole and posterior cingulate regions. The identified insula region exhibited a strong functional interaction with the salience network (including the bilateral insula and the dorsal ACC, dACC) while the vACC coupled strongly with core regions of the default mode network (DMN), including the medial prefrontal cortex, posterior cingulate and the bilateral angular gyrus while the STG showed strong functional interaction with the bilateral temporal regions (**Fig. 3a-d**).

**Fig. 3.**
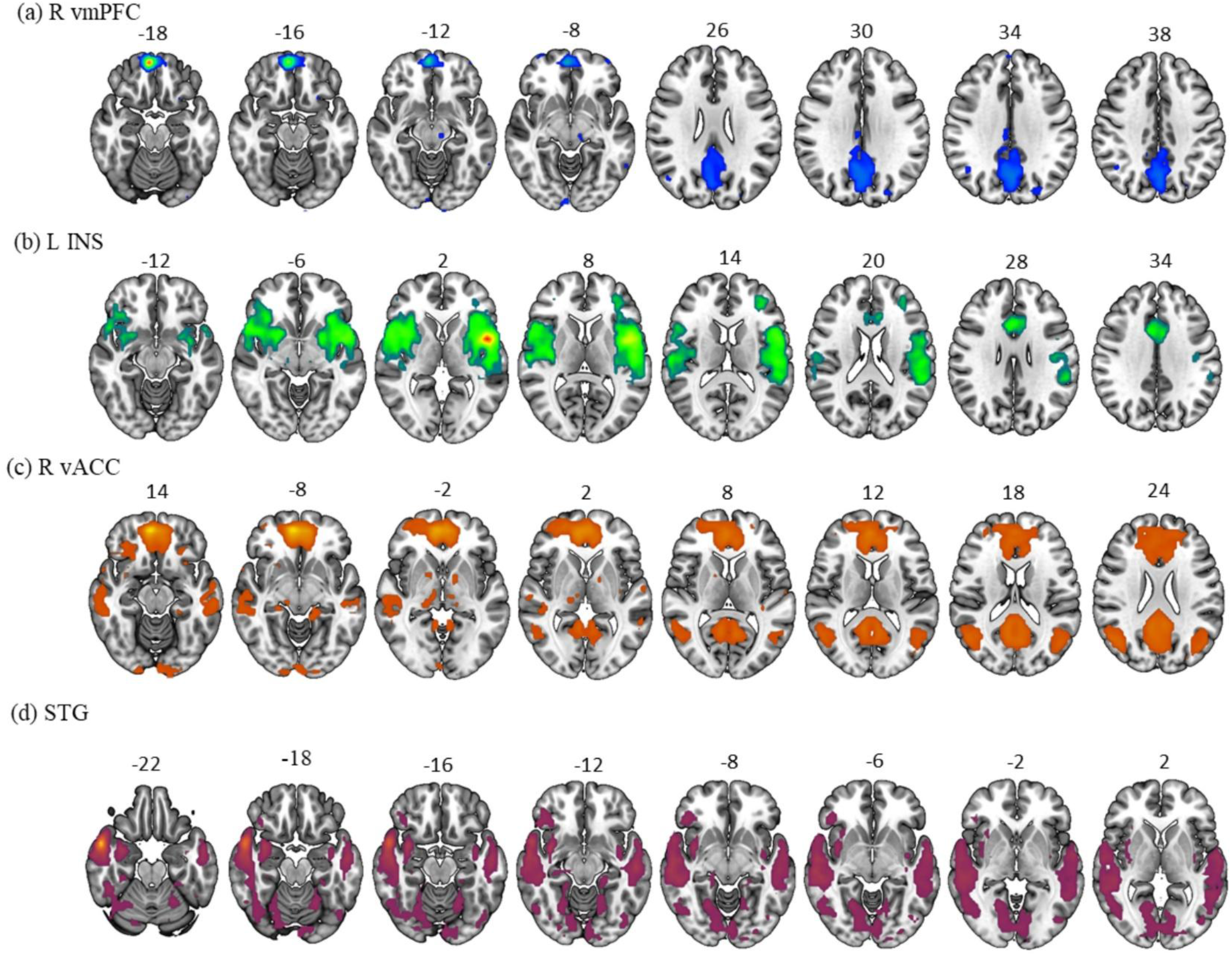
Seed-based functional connectivity of the common regions of **a** ventromedial prefrontal cortex, **b** insula, **c** ventral anterior cingulate cortex and **d** superior temporal gyrus.

Behavioral-level analyses revealed that the vmPFC was associated with social and theory of mind related functions. The insula strongly associated with pain, motor and primary functions (coordination and regulation of body functions) while the STG showed stronger associations with theory of mind, social and autobiographical functions. The vACC coupled more with the default network, autobiographical and theory of mind related functions (**Fig. 4a-b**).

**Fig. 4.**
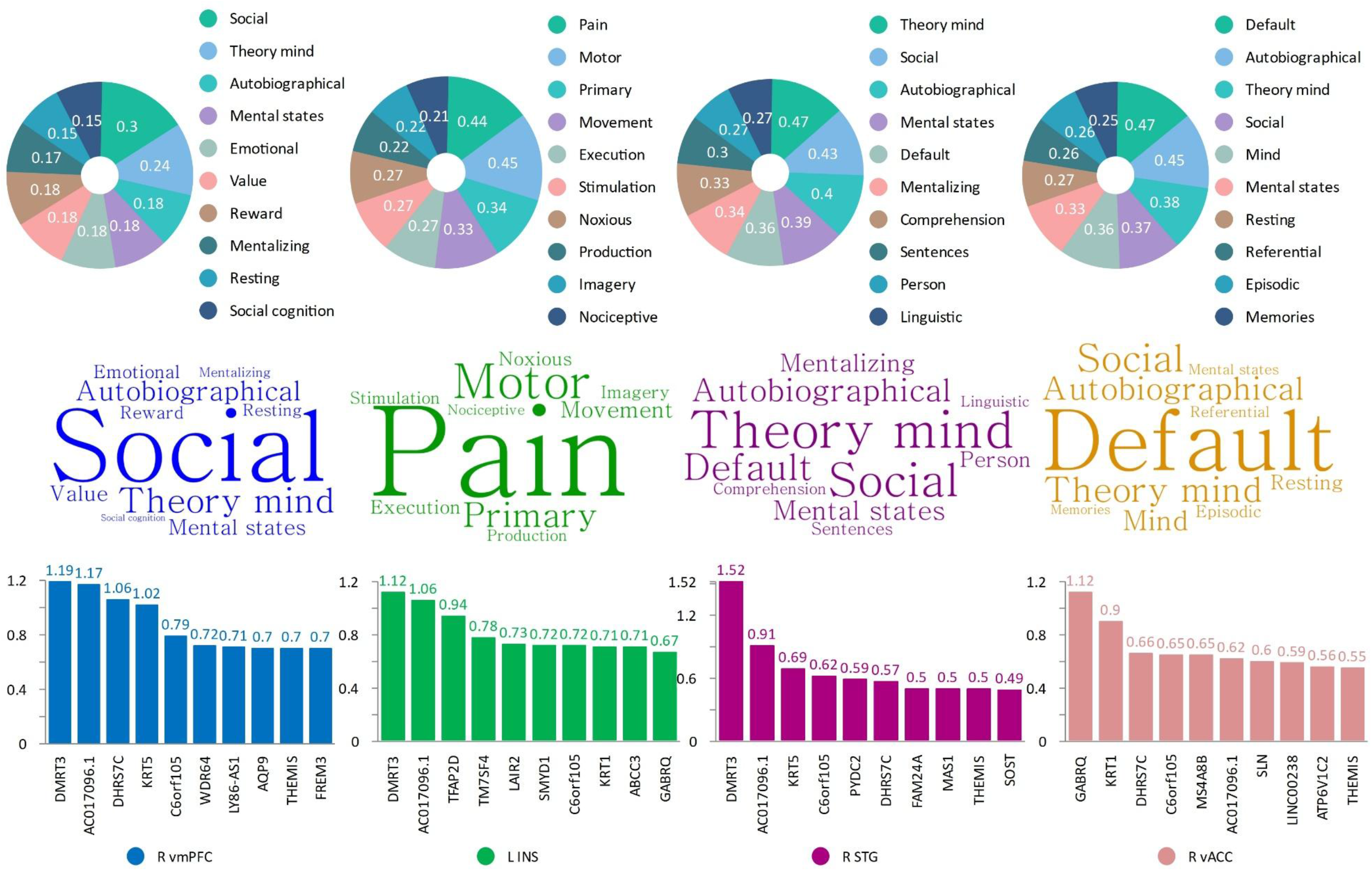
Panel **a** displays the top ten behavioral terms of the common regions of gray matter decreases of the vmPFC, insula, STG and ventral anterior cingulate cortex [respectively, left-right]. Panel **b** is a word cloud display of the behavioral terms listed in panel a. Panel **c** shows the top ten genetic annotations of the common regions.

Genetic-level characterization showed high gene expressions of the DMRT3 gene in the vmPFC, insula and STG. DMRT3 gene is mostly responsible for nerve impulse transmissions. However, the vACC revealed high expression of GABRQ gene which is responsible for mediating responses in the central nervous system (**Fig. 4c**). Receptor-level analysis revealed overall higher dopamine (DAT) and serotonin (5HT_1b_) mean values in the STG and insula respectively. Specifically, mean densities of the vmPFC were 0.0398 and 0.1365 for DAT and 5HT_1b_ respectively (t=-11.95, df=1028, *SD*=0.1298). The insula had mean densities of 0.1309 and 0.1730 for DAT and 5HT_1b_ respectively (t=-4.45, df=1028, *SD*=0.1519). The STG had mean densities of 0.1348 and 0.1319 for DAT and 5HT_1b_ respectively (t=0.44, df=1028, *SD*=0.10). The vACC had mean densities of 0.1129 and 0.1087 for DAT and 5HT_1b_ respectively (t=0.57, df=1028, *SD*=0.11). More details are presented in **Supplementary Fig. 5a.**

### Tests for sensitivity, heterogeneity and publication bias

The jackknife sensitivity analyses confirmed the robustness of the meta-analysis findings. There was low to moderate levels of heterogeneity across all meta-analyses. Publication bias did not reach statistical significance as shown by the p-values and funnel plots in the supplements. Individual results are shown in Table 1.

### Exploratory analyses

Overall, the positive and negative associations revealed by the meta-regression analyses on number of females and age were not statistically significant across the three disorders (**Figure 5b-d).** Exploratory linear model analysis did not reveal significant medication effects in SZ and MDD and supplementary meta-analysis of non-medication studies validated the stability of the original findings (**Supplementary Fig. 3**)

## Discussion

The present meta-analysis is to our knowledge the first to systematically determine transdiagnostic GMV decreases in three common psychiatric disorders with high co-morbidity and symptomatic overlap in the domains of anhedonia, executive and social functioning. Meta-analytic characterization of SZ, MDD and AUD revealed: a) shared GMV decreases in the ventromedial prefrontal cortex (vmPFC) extending to the ventral anterior cingulate cortex (vACC), insula and superior temporal gyrus (STG) reflecting transdiagnostic deficits, b) robust GMV decreases within each disorder, with comparably focused decreases in MDD, while SZ and AUD exhibited more widespread decreases, c) regions exhibiting shared GMV decreases were characterized by functional interactions with cortical midline structures and default mode core regions (vmPFC, vACC), the salience network (INS), and bilateral temporal regions (STG) and behavioral domains centered around social cognitive and autobiographical functions. Additional analyses highlighted the involvement of dopaminergic and serotonergic neurotransmitters as well as genetic annotations associated with architecture and function of the nervous system. Exploratory meta-regression suggested no significant associations between age and gender (number of females) across the main meta-analytic findings. The identified regions may mediate shared symptomatic deficits in reward/hedonic, executive and social functioning, with further meta-analytic characterization supporting a neurobiologically-informed account of transdiagnostic social and autobiographical deficits.

### Neurostructural assessment of transdiagnostic GMV abnormalities across disorders

Capitalizing on data from over 13,000 individuals allowed us to robustly determine transdiagnostic reductions in GMV in the vmPFC extending to the vACC, insula and STG. The vmPFC has been identified as a key region linked with the psychopathology of several psychiatric disorders across several meta-analyses [194]. Dense projections of the vmPFC with the hypothalamus, amygdala and ventral striatum mediate its critical role in emotion regulation and fear as well as reward and motivational processes [195, 196]. Animal and human studies have consistently demonstrated a role of the vmPFC in representation and updating of reward values and outcomes as well as social cognitive functions [197–199].

Previous studies suggested a link between vmPFC dysfunction and value and decision-making deficits in AUD, with SZ and MDD showing shared alterations in value, decision-making and social functions related to the vmPFC [200]. Severity of depressive symptoms and associated cognitive and social impairments were associated with variations in the vmPFC in depression [194, 201]. Previous meta-analyses have found significantly reduced GMV in medial prefrontal regions across mood but also anxiety disorders [13].

With respect to the further functional network characterization of this region we observed strong coupling with cortical midline regions of the core DMN and meta-analytic associations with social domains, including theory of mind, autobiographic memory or mentalizing, suggesting that alterations in this region may contribute to transdiagnostic social deficits [194]. The observed alterations in the vmpFC may thus contribute to social impairments across disorders including altered empathy and processing of social values [35, 202]. Further analyses revealed that monoamine neurotransmitters involved in reward and social processes, specifically dopamine and serotonin exhibited a high expression in this region. The literature has extensively unraveled the core roles of dopaminergic and serotonergic signaling and how implications of these signals underlie neuropsychiatric disorders. Impaired serotonergic signaling in the vmPFC have been described in animal models and this has specifically implicated vmPFC stimulation [203]. Taken together, the results from GMV loss, functional networks, behavioral and molecular characterizations may suggest several possibilities suggesting that vmPFC deficits reflect impaired a) regulation of negative emotions, b) reward and value representation, or c) social and autobiographical functioning in patients with SZ, MDD and AUD.

Alterations of the insula (both anterior and posterior) have been identified as a common psychopathological substrate in several neuropsychiatric disorders [41, 204]. On the anatomical and functional level, recent models have proposed that insula alterations may be implicated in depressive, schizophrenic and substance use disorders [205–207]. Anatomical alterations of the insula have been strongly associated with negative symptoms in SZ, with significant GMV reductions observed in several studies [208] suggesting associations with social and cognitive deficits. In MDD, SZ and AUD, neuroanatomical abnormalities of the insula have been frequently reported [10, 89, 209]. Insula alterations have been involved in several domains, including alterations in incentive motivation, interoception, social functioning and pain [210–212]. Our behavioral characterization revealed that the identified insula region was associated with pain, in line with evidence from the literature indicating its crucial role in altered pain and pain empathic processing especially in MDD [213] and SZ [214]. Of the four common regions identified in the three disorders, the insula recorded the highest mean value of serotonin (5HT_1B_) neurotransmitter. The serotonin system is associated with mood and social interactions in humans, which may link it to anhedonia and social deficits across psychiatric disorders [215]. On the network level, the insula coupled strongly with regions encompassing the salience network which help integrate sensory information and also help in assessing the salience of stimuli, therefore, structural decreases in the region may cause challenges in identifying important social stimuli and responding to emotional states.

The ventral ACC (vACC) is associated with several cognitive and emotional processes and is often described as an interconnection of these two processes [216, 217]. GMV decreases in the vACC has been associated with the negative symptoms including anhedonia and emotional dysregulations in psychiatric disorders, with a recent meta-analysis also suggesting robust GMV ACC decreases in AUD and post traumatic disorders [19]. Mirroring the characterization of the adjacent vmPFC this region exhibited strong connections with the core DMN and a behavioral characterization including social and autobiographical domains. Together with the vACC’s role in emotional conflict and regulation, this may indicate that dysfunctions in this region may underlie impaired integration of affective regulation with social and autobiographical processes.

Dysfunctions in social and autobiographical domains are further supported by our observation of robust decreases in the STG GMV across the disorders. This region plays a crucial role in social cognition and integration of affective and mnemonic processes strongly involved in the pathophysiology of several psychiatric disorders. While previous case control studies and meta-analyses on morphometric alterations in this region in these disorders remained inconsistent [21, 218], a growing number of studies has implicated dysfunctions in this region with deficits in social functioning [219].

Strong network connections of the identified STG region with the extended DMN including anterior temporal regions and the behavioral characterization further confirmed an involvement in social and autobiographical processing. Impairments in the STG may mediate deficits in these domains which have been consistently been identified as factors contributing to the development and maintenance of psychiatric disorders [220].

While MDD, SZ, and AUD exhibit overlapping deficits in the domains of reward processing/anhedonia, executive functions, and social processes, a neurobiological characterization of the regions showing decreased GMV integrity across these disorders supports a neurobiologically informed framework indicating transdiagnostic dysfunctions in the domains of autobiographical and social processing.

## Limitations

There are some limitations to be considered in the present meta-analysis. First, SZ and MDD studies included in the main analyses had partly different medication with the literature suggesting that patients exposed to medication may show progressive structural changes in GMV [221, 222]. However, our sub-group meta-analyses focusing on studies with unmediated samples confirmed the main findings (Supplementary Fig. 3). Secondly, information on duration of duration of illness or alcohol use was not extensively provided across the three disorders. This important variable may affect the findings if the differences are extreme across studies and this should be further explored in future meta-analyses. Thirdly, we performed a coordinate-based meta-analysis which are more generalized and may often be less accurate than image-based meta-analyses. Sharing of individual level data in future studies may help to overcome this limitation. Fourth, some studies did not provide information regarding age of onset of illness. It is recommended that original studies should provide detailed clinical information of all included patients to ensure more comprehensive analyses in the future.

## Conclusion

Taken together, we found specific GMV abnormalities across several cortical and subcortical brain regions supporting a neurobiologically-informed model of transdiagnostic dysregulations in social and autobiographical domains. Given the robust transdiagnostic deficits in GMV alterations in the vmPFC, insula, ventral ACC and superior temporal gyrus, transdiagnostic interventions may target these regions and/or social and autobiographical deficits.

## Author contributions

MCB, BKB, BB conceived and designed the study. MCB, JH, MH, ATH, XL, JZ, BKB, BB acquisition, analyzed and interpreted the data. MCB, BKB, BB drafted the original manuscript. JZ, KK, XH and JY provided critical conceptual and intellectual input. MCB, JH, and BKB performed statistical analysis. BB obtained funding. MH, ATH, XL, BKB and BB provided administrative and technical support. BKB and BB provided supervision.

## Competing interests

The authors declare no competing interests.

## Funding/Support

This study was supported by the Health and Medical Research Fund (HMRF, 23243971), Ministry of Science and Technology, MOST2030, Grant No. 2022ZD0208500 and a seed grant for collaborative research from The University of Hong Kong (HKU 2407102536).

## Role of the Funder/Sponsor

The funding sources had no role in the design and conduct of the study; collection, management, analysis, and interpretation of the data; preparation, review, or approval of the manuscript; and the decision to submit the manuscript for publication. Any opinions, findings, conclusions, or recommendations expressed in this publication do not reflect the views of the Government of the Hong Kong Special Administrative Region or the Innovation and Technology Commission.

## Data availability

The studies included in the meta-analysis have been cited in the manuscript. Coordinates will be made available from the corresponding author upon request.

## Notes

### Competing Interest Statement

The authors have declared no competing interest.

### Summary of Updates

Version includes additional technical specifications.

