## Supplementary Information for "Transdiagnostic gray matter volume reductions implicating social dysfunction in depression, schizophrenia, and alcohol use disorder: A neuroimaging meta-analysis"

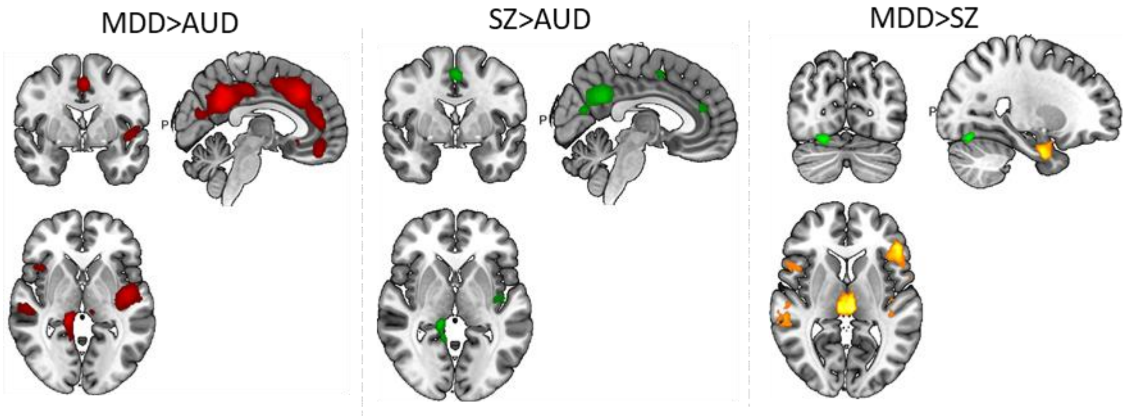


**Supplementary Fig. 1.** Disorder-specific gray matter alterations in Major depressive disorder, Schizophrenia and Alcohol Use Disorder

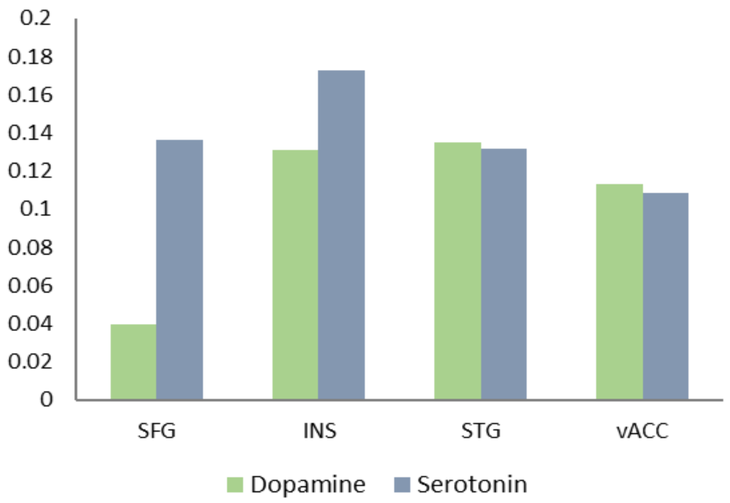


**Supplementary Fig. 2.** Receptor level comparison of DAT and $5HT_{1B}$ in the common regions of decreased gray matter in the three disorders**.**

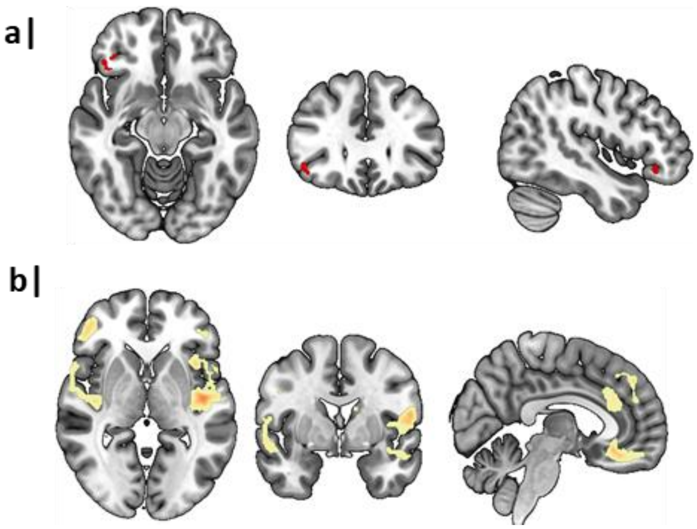


**Supplementary Fig. 3.** Meta-analysis results of unmedicated studies in MDD (a) and SZ (b).

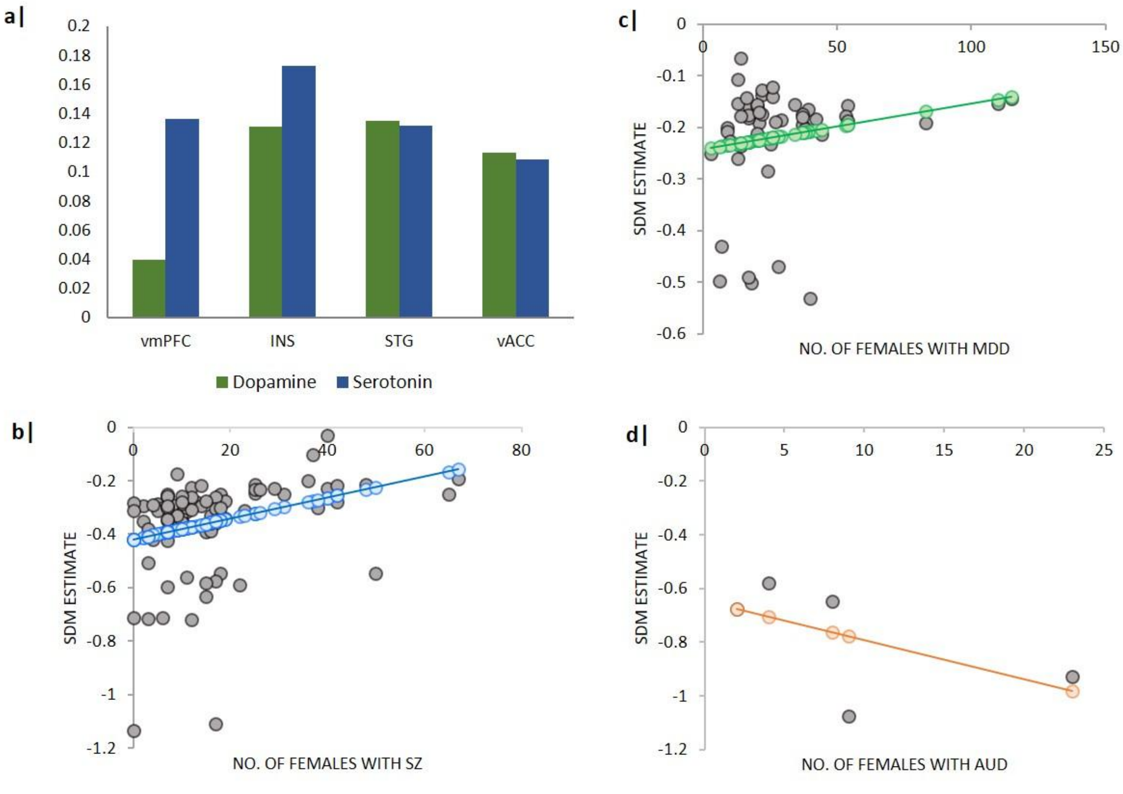


**Supplementary Fig. 4.** Receptor analyses of the common regions and meta-regression analyses showing effects of gender in the main analyses. **a** Comparison of the mean values of dopamine and serotonin in the common regions, **b** Subsequent analyses for effects of gender (number of female patients) across the three disorders.

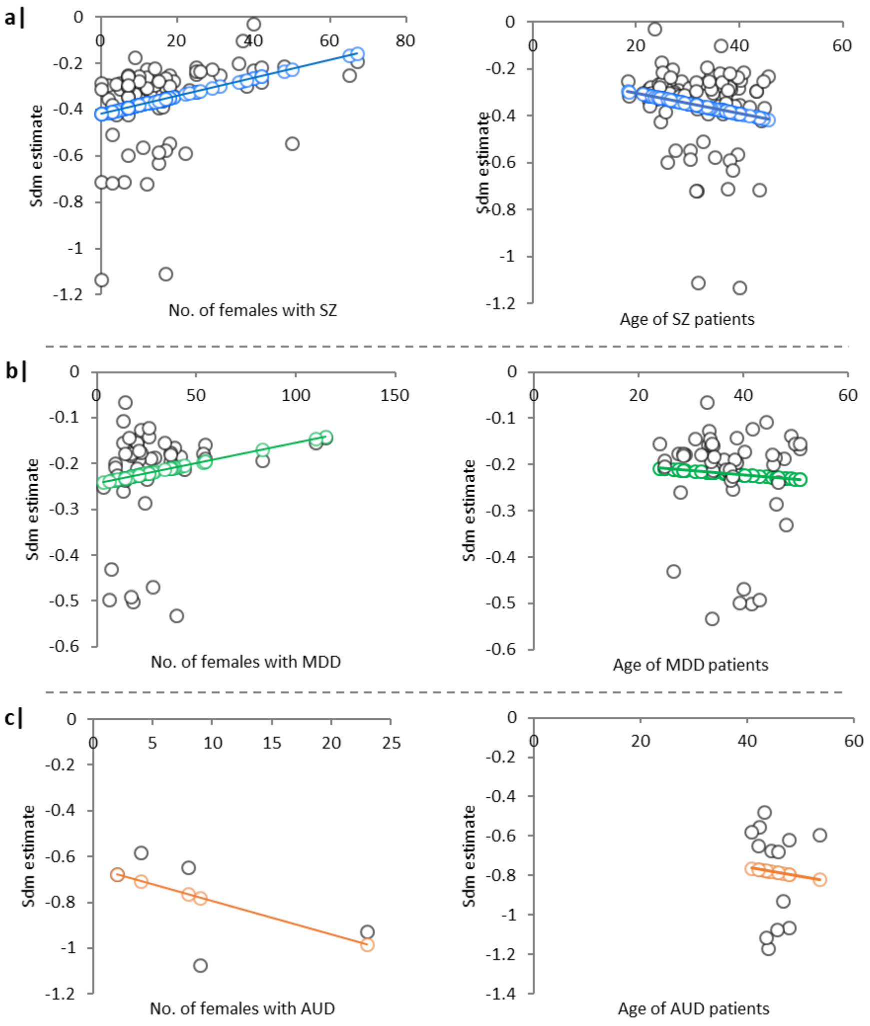


**Supplementary Fig. 5.** Comparison of the regression analysis of age and number of females in SZ (a), MDD (b) and AUD (c) in adults.

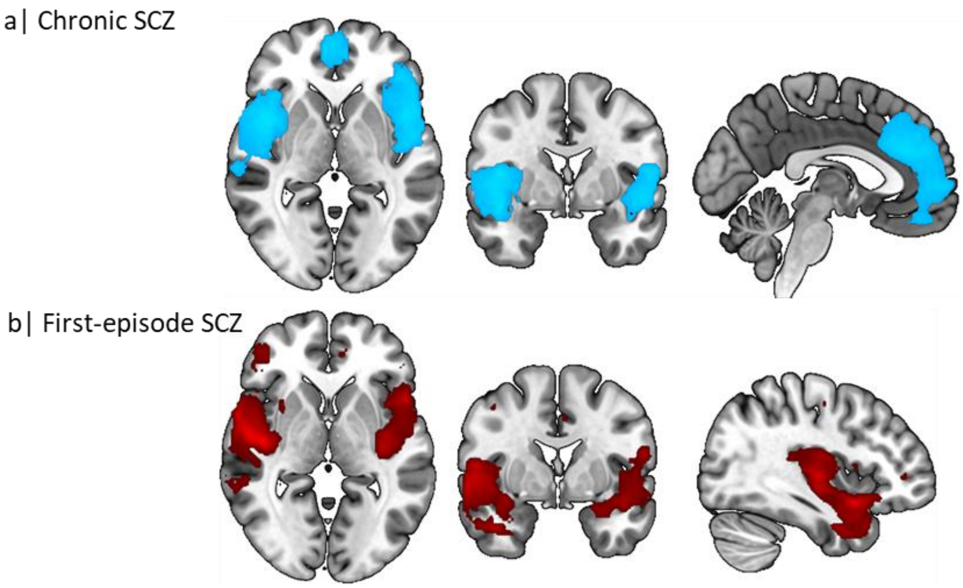


**Supplementary Fig. 6.** Comparison between First-episode SZ and Chronic SZ

**Supplementary Table 1.** PRISMA checklist 2020

| Section and topic | Item # | Checklist item | Reported on page # |
|---|---|---|---|
| **TITLE** | | | |
| Title | 1 | Identify the report as a systematic review. | 1 |
| **ABSTRACT** | | | |
| Abstract | 2 | See the PRISMA 2020 for Abstracts checklist | 3 |
| **INTRODUCTION** | | | |
| Rationale | 3 | Describe the rationale for the review in the context of existing knowledge. | 4,5,6,7 |
| Objectives | 4 | Provide an explicit statement of the objective(s) or question(s) the review addresses. | 4,5,6,7 |
| **METHODS** | | | |
| Eligibility criteria | 5 | Specify the inclusion and exclusion criteria for the review and how meta-analyses were grouped for the synthesis. | 8 |
| Information sources | 6 | Specify all databases, registers, websites, organisations, reference lists and other sources searched or consulted to identify studies. Specify the date when each source was last searched or consulted. | 7,8 |

| Section and topic | Item # | Checklist item | Reported on page # |
|---|---|---|---|
| Search strategy | 7 | Present the full search strategies for all databases, registers and websites, including any filters and limits used. | 7,8 |
| Selection process | 8 | Specify the methods used to decide whether a meta-analysis met the inclusion criteria of the review, including how many reviewers screened each record and each report retrieved, whether they worked independently, and if applicable, details of automation tools used in the process. | 8 |
| Data collection process | 9 | Specify the methods used to collect data from reports, including how many reviewers collected data from each report, whether they worked independently, any processes for obtaining or confirming data from study investigators, and if applicable, details of automation tools used in the process | 8 |
| Data items | 10a | List and define all outcomes for which data were sought. Specify whether all results that were compatible with each outcome domain in each study were sought (e.g. for all measures, time | - |

| Section and topic | Item # | Checklist item | Reported on page # |
|---|---|---|---|
| | | points, analyses), and if not, the methods used to decide which results to collect. | |
| | 10b | List and define all other variables for which data were sought (e.g. participant and intervention characteristics, funding sources). Describe any assumptions made about any missing or unclear information. | Supplementary Table 2,3,4 |
| Meta-analysis quality assessment | 11 | Specify the methods used to assess quality in the included meta-analyses, including details of the tool(s) used, how many reviewers assessed each meta-analysis and whether they worked independently, and if applicable, details of automation tools used in the process | 8,9 |
| Effect measures | 12 | Specify for each outcome the effect measure(s) (e.g. risk ratio, mean difference) used in the synthesis or presentation of results. | 8,9 |
| Synthesis methods | 13a | Describe the processes used to decide which meta-analyses were eligible for each synthesis (e.g. tabulating the study intervention | 9, Supplementary Table 2,3,4 |

| Section and topic | Item # | Checklist item | Reported on page # |
|---|---|---|---|
| | | characteristics and comparing against the planned groups for each synthesis. | |
| | 13b | Describe any methods required to prepare the data for presentation or synthesis, such as handling of missing summary statistics, or data conversions. | 9 |
| | 13c | Describe any methods used to tabulate or visually display results of individual meta-analyses. | 9 |
| | 13d | Describe any methods used to synthesise results and provide a rationale for the choice(s). | 8,9,10,11 |
| | 13e | Describe any methods used to explore possible causes of heterogeneity among study results (e.g. subgroup analysis, meta-regression). | 12 |
| | 13f | Describe any sensitivity analyses conducted to assess robustness of the synthesised results | 12 |
| Reporting bias assessment | 14 | Describe any methods used to assess risk of bias due to missing results in a synthesis (arising from reporting biases). | 12 |
| Certainty | 15 | Describe any methods used to assess certainty | - |

| Section and topic | Item # | Checklist item | Reported on page # |
|---|---|---|---|
| assessment | | (or confidence) in the body of evidence for an outcome. | |
| **RESULTS** | | | |
| Study selection | 16a | Describe the results of the search and selection process, from the number of records identified in the search to the number of studies included in the review, ideally using a flow diagram. | Figure 1 |
| | 16b | Cite studies that might appear to meet the inclusion criteria, but which were excluded, and explain why they were excluded. | Figure 1 |
| Study characteristics | 17 | Cite each included study and present its characteristics. | 12,13 Supplementary Table 2,3,4 |
| Quality of meta-analyses | 18 | Present assessments of quality for each included meta-analysis. | 15 |
| Results of individual meta-analyses | 19 | For all outcomes, present, for each meta-analysis: (a) summary statistics for each group (where appropriate) and (b) an effect estimate and its precision (e.g. confidence/credible interval), ideally using structured tables or plots. | 13, Table 1, Figure 2 |

| Section and topic | Item # | Checklist item | Reported on page # |
|---|---|---|---|
| Results of syntheses | 20a | For each synthesis, briefly summarise the characteristics and quality among contributing meta-analyses. | 13,15 |
| | 20b | Present results of all statistical syntheses conducted. If meta-analysis was done, present for each the summary estimate and its precision (e.g. confidence/credible interval) and measures of statistical heterogeneity. If comparing groups, describe the direction of the effect. | 13,15, Table 1 |
| | 20c | Present results of all investigations of possible causes of heterogeneity among study results. | 15, Table 1 |
| | 20d | Present results of all sensitivity analyses conducted to assess the robustness of the synthesised results. | 13,15, Table 1 |
| Reporting biases | 21 | Present assessments of risk of bias due to missing results (arising from reporting biases) for each synthesis assessed. | 15, Table 1 |
| Certainty of evidence | 22 | Present assessments of certainty (or confidence) in the body of evidence for each outcome assessed. | - |

| Section and topic | Item # | Checklist item | Reported on page # |
|---|---|---|---|
| **DISCUSSION** | | | |
| Discussion | 23a | Provide a general interpretation of the results in the context of other evidence | 16,17,18,19,20 |
| | 23b | Discuss any limitations of the evidence included in the review. | 20,21 |
| | 23c | Discuss any limitations of the review processes used. | 20,21 |
| | 23d | Discuss implications of the results for practice, policy, and future research | 21 |
| **OTHER INFORMATION** | | | |
| Registration and protocol | 24a | Provide registration information for the review, including register name and registration number, or state that the review was not registered. | 7 |
| | 24b | Indicate where the review protocol can be accessed, or state that a protocol was not prepared. | 7 |
| | 24c | Describe and explain any amendments to information provided at registration or in the protocol. | 7 |
| Support | 25 | Describe sources of financial or non-financial | 22 |

| Section and topic | Item # | Checklist item | Reported on page # |
| --- | --- | --- | --- |
| | | support for the review, and the role of the funders or sponsors in the review. | |
| Competing interests | 26 | Declare any competing interests of review authors. | 22 |
| Availability of data, code, and other materials | 27 | Report which of the following are publicly available and where they can be found: template data collection forms; data extracted from included studies; data used for all analyses; analytic code; any other materials used in the review. | 22 |

**Supplementary Table 2.** Demographic characteristics of patients with MDD

| Study | No. of patients(F) | No. of HCs (F) | Mean age of patients (SD) | Mean age of HCs (SD) | Duration of illness | Medication |
|---|---|---|---|---|---|---|
| Wang 2020 [4] | 36 (20) | 27 (14) | 34.11 (10.39) | 32.44 (8.57) | 10.19 | X |
| Zhang 2021 [5] | 20 (13) | 20 (10) | 28 (9.1) | 31.7 (11.4) | NA | X |
| Mortel 2022 [6] | 88 (39) | 27 (11) | 50.77 (12) | 60.7 (8) | NA | √ |
| Li 2017 [7] | 23 (20) | 25 (20) | 49.91 (8.44) | 49.2 (10.25) | NA | X |
| Zhang 2020 [8] | 30 (21) | 63 (39) | 25 (NA) | 23 (NA) | NA | X |
| Yang 2017 [9] | 82 (53) | 82 (53) | 28.84 (9.17) | 27.72 (8) | 59.85 | X |
| Meng 2020 [10] | 159 (83) | 53 (25) | 33.71 (10.21) | 35.64 (8.66) | NA | X |
| Romanczuk 2014 [11] | 63 (38) | 63 (38) | 25 (NA) | 26 (NA) | NA | X |
| Liu 2012 [12] | 17 (7) | 17 (7) | 26.71 (7.73) | 24.24 (4.41) | 2.59 | √ |
| Lan 2016 [13] | 27 (18) | 27 (15) | 41.5 (16) | 39.2 (16.7) | NA | NA |
| Brosch 2022 [14] | 110 (54) | 110 (56) | 38.35 (11.7) | 38.83 (12.21) | NA | √ |
| Depping 2015 [15] | 22 | 22 | 33.5 (8.9) | 31.4 (11.2) | 5.5 | √ |
| Guo 2014 [16] | 44 (22) | 44 (24) | 27.52 (8.57) | 29.39 (6.7) | 19.61 | X |
| Jung 2014 [17] | 24 (17) | 29 (21) | 43 (10.1) | 43.6 (13.4) | 33.3 | √ |
| Kong 2014 [18] | 28 (17) | 28 (14) | 34.42 (8.24) | 32.07 (9.27) | 2.11 | X |
| Lai & Wu 2014 [19] | 38 (20) | 27 (15) | 36.57 (5.46) | 38.29 (11.8) | 4.68 | X |
| Lai & Wu 2015 [20] | 53 (28) | 54 (29) | 40.07 (8.99) | 40.38 (10.51) | 5.03 | X |
| Lai 2016 [21] | 15 (9) | 27 (15) | 37.46 (5.93) | 38.29 (11.8) | 4.2 | √ |
| Liu 2022 [22] | 64 (37) | 61 (25) | 28.47 (9.74) | 30.49 (9.55) | 15.2 | X |
| Mak 2009 [23] | 17 | 17 | 45.5 (8.5) | 45.8 (9.8) | NA | √ |
| Nan 2020 [24] | 166 (115) | 166 (115) | 33.69 (9.99) | 34.64 (10.98) | 11.33 | X |
| Opel 2016 [25] | 20 (10) | 20 (10) | 37.9 (10.9) | 36.3 (12.1) | 139.4 | X |
| Ozalay 2016 [26] | 24 | 24 | 46.2 (3.9) | 47.3 (5.6) | 13.3 | X |
| Peng 2011 [27] | 22 (14) | 30 (19) | 46.7 (8.9) | 45.9 (9) | 8.6 | √ |
| Xu 2019 [28] | 11 (6) | 12 (6) | 39.27 (7.84) | 39.08 (7.4) | 25.81 | X |
| Zhuo 2017 [29] | 45 (26) | 48 (27) | 38.8 (13.3) | 38.6 (10.5) | 36.5 | √ |
| Chen 2018 [30] | 36 (16) | 47 (25) | 30.7 (8.5) | 29.7 (9.2) | 32.1 | X |
| Abe 2010 [31] | 21 (10) | 42 (20) | 48.1 (13.5) | 48 (13.2) | 14.17 | √ |
| Grieve 2013 [32] | 102 (54) | 34 (16) | 33.8 (13.1) | 31.5 (12.4) | 135.6 | X |
| Kandilarova 2019 [33] | 39 (29) | 42 (29) | 47.7 (13.9) | 42.6 (13.7) | 129.6 | √ |
| Klauser 2015 [34] | 56 (40) | 33 (21) | 34.02 (8.96) | 34.71 (9.93) | 123.48 | √ |
| Lee 2011 [35] | 47 (42) | 51 (45) | 46 (9.1) | 45.7 (8) | 46.7 | √ |
| Lu 2018 [36] | 76 (44) | 86 (43) | 34.27 (8.37) | 33.35 (7.62) | 46.75 | X |
| Lu 2019 [37] | 30 (13) | 48 (30) | 23.98 (5.25) | 21.5 (3.84) | 30.1 | X |

| | | | | | | |
|---|---|---|---|---|---|---|
| Nakano 2014 [38] | 36 (22) | 54 (27) | 49 (11.4) | 45.4 (16.1) | 66.7 | √ |
| Salvadore 2011 [39] | 58 (37) | 107 (60) | 38.8 (11.1) | 36.2 (10.3) | 220.8 | √ |
| Ueda 2016 [40] | 30 (13) | 48 (13) | 44.3 (13) | 41.2 (11.4) | NA | X |
| Wagner 2011 [41] | 30 (25) | 30 (25) | 37.55 (11.5) | 35.1 (10.4) | 71.4 | X |
| Chen 2016 [42] | 27 (14) | 28 (14) | 33 (10.8) | 33 (11.7) | 79 | X |
| Bergouignan 2009 [43] | 20 (17) | 21 (14) | 33.16 (9.58) | 28.21 (5.5) | 101.4 | √ |
| Scheuerecker [44] | 13 (3) | 15 (5) | 37.9 (10.1) | 35.5 (10.9) | 52.3 | X |
| Arnone 2013 [45] | 39 (27) | 66 (46) | 36.3 (8.8) | 32.1 (9.3) | NA | X |
| Sprengelmeyer 2011 [46] | 17 (9) | 21 (12) | 45.6 (12.3) | 42 (12.9) | NA | √ |
| ChenS 2024 [47] | 163 (110) | 134 (35) | 34 (12) | 36 (10) | 72 | X |
| Borgers 2024 [48] | 50 (26) | 21 (11) | 41.79 (11.32) | 40.57 (12.98) | NA | √ |

Abbreviations: HC, healthy controls, NA, not available, No. number, SD, standard deviation

**Supplementary Table 3.** Demographic characteristics of patients with SZ

| Study | Patients | Healthy controls | Mean age of patients (s.d) | Mean age of HCs (s.d) | Duration of illness | Medication |
|---|---|---|---|---|---|---|
| Amann 2016 [49] | 45 (19) | 45 (19) | 43.2 (9.1) | 43.3 (9.9) | 258 | √ |
| Ananth 2002 [50] | 20 (10) | 20 (10) | 37.8 (9.5) | 38.6 (9.7) | 184.2 | √ |
| Asami 2012 [51] | 33 (5) | 36 (6) | 22.5 (6.7) | 22.9 (3.8) | 4.9 | √ |
| Bassitt 2007 [52] | 50 (12) | 30 (9) | 31.7 (7.1) | 31.2 (7.6) | 136.8 | √ |
| Bonilha 2008 [53] | 14 (2) | 13 (NA) | 40 (NA) | 35 (NA) | NA | X |
| Borgwardt 2010 [54] | 28 (6) | 34 (10) | 37.7 (9.1) | 39.3 (9.5) | 315.6 | √ |
| Brown 2011 [55] | 17 (9) | 21 (11) | 44.8 (6.8) | 45 (10.2) | 229.2 | √ |
| Cascella 2010 [56] | 50 (13) | 90 (47) | 39.8 (11.1) | 46.3 (12.7) | 189 | √ |
| Chang 2016 [57] | 60 (31) | 71 (44) | 18.3 (3.09) | 20.6 (3.48) | 8.1 | √ |
| Chang 2020 [58] | 51 (18) | 128 (68) | 28.73 (9.71) | 33.34 (11.85) | 58.1 | X |
| Chua 2007 [59] | 26 (17) | 38 (22) | 32 (10) | 33 (8.1) | 4 | X |
| Cooke 2008 [60] | 52 (12) | 30 (6) | 38.35 (9.89) | 32.13 (12.38) | 166.8 | √ |
| Corradi 2012 [61] | 68 (25) | 77 (37) | 40.15 (12.06) | 39.63 (10.73) | 115.9 | √ |
| Cui 2011 [62] | 23 (7) | 36 (15) | 24.78 (5.09) | 29.56 (6.7) | 48.5 | √ |
| de Castro 2011 [63] | 28 (11) | 20 (9) | 18.6 (4.9) | 20.5 (5.7) | 7.1 | √ |
| Delvecchio 2017 [64] | 61 (25) | 59 (24) | 40.8 (10.8) | 40.2 (11.3) | 175.2 | √ |
| Euler 2009 [65] | 19 (2) | 23 (7) | 43.26 (10.5) | 43.3 (11.9) | NA | √ |

| | | | | | | |
|---|---|---|---|---|---|---|
| Filippi 2014 [66] | 43 (19) | 17 (11) | 29.3 (7.4) | 30.7 (8.6) | 7.9 | Χ |
| Fukuta 2013 [67] | 40 | 50 | 45.6 (8.1) | 45 (8.7) | 276 | √ |
| Guo 2014 [68] | 19 (16) | 19 (14) | 24 (7.49) | 25.2 (8.07) | 9.4 | √ |
| Guo 2014 [69] | 51 (18) | 41 (17) | 22.5 (4.1) | 22.8 (3.9) | 8.4 | √ |
| Guo 2014 [70] | 49 (19) | 50 (27) | 22.7 (4.62) | 23.5 (2.49) | 22.5 | x |
| Guo 2019 [71] | 33 (17) | 33 (17) | 24.3 (8.8) | 23.8 (8.4) | NA | x |
| Herold 2009 [72] | 18 (7) | 21 (10) | 28.7 (10.3) | 27.4 (6.5) | 40.8 | √ |
| Hirao 2008 [73] | 20 (10) | 20 (10) | 36.7 (7.6) | 35 (7.1) | 127.2 | √ |
| Honea 2008 [74] | 169 (37) | 212 (109) | 36.4 (9.46) | 33.3 (9.86) | NA | √ |
| Hooker 2011 [75] | 21 (4) | 17 (4) | 44.3 (10.18) | 43.8 (11.75) | 293.6 | √ |
| Hu 2013 [76] | 51 (17) | 59 (21) | 22.3 (3.95) | 23.2 (2.58) | 11.1 | √ |
| Huang 2015 [77] | 36 (17) | 18 (9) | 22.65 (5.29) | 25.1 (2.44) | 18.3 | Χ |
| Jayakumar 2005 [78] | 18 (9) | 18 (9) | 24.9 (6.3) | 25.7 (7.5) | 10.3 | Χ |
| Job 2002 [79] | 34 (11) | 36 (19) | 21.35 (3.66) | 21.17 (2.37) | NA | √ |
| Kawada 2009 [80] | 26 (15) | 26 (15) | 36.7 (8.6) | 36.3 (8.8) | 118.8 | √ |
| Kim 2017 [81] | 22 (10) | 22 (10) | 31.7 (10.1) | 31.6 (9.5) | 110.4 | √ |
| Koelkebeck 2013 [82] | 18 (6) | 30 (16) | 34.9 (10.1) | 39.1 (10.8) | 141.6 | √ |
| Lee 2020 [83] | 65 (29) | 65 (37) | 37 (7.88) | 34.5 (8.93) | 186.2 | √ |
| Lei 2015 [84] | 44 (18) | 44 (18) | 22.9 (6.89) | 22.6 (6.25) | 19.9 | √ |

| | | | | | | |
|---|---|---|---|---|---|---|
| Li 2020 [85] | 86 (40) | 86 (41) | 23.5 (6.9) | 24 (6.3) | 14.2 | √ |
| Liao 2015 [86] | 93 (36) | 99 (46) | 27 (6.6) | 25.8 (5.4) | 54.1 | √ |
| Lui 2009 [87] | 68 (38) | 68 (37) | 24.2 (8.6) | 24.7 (8.8) | 8.6 | Χ |
| Mane 2009 [88] | 15 (3) | 11 (3) | 25.6 (5.77) | 30.3 (4.36) | 5.8 | √ |
| Martin 2014 [89] | 21 (65) | 10 (28) | 44.6 (10.7) | 46.5 (9.6) | NA | √ |
| Molina 2010 [90] | 30 (10) | 40 (17) | 25.8 (5) | 29.4 (9) | 14.5 | Χ |
| Molina 2011 [91] | 38 (12) | 24 (8) | 34.4 (10.5) | 34.6 (8.6) | 160.8 | √ |
| Nagashima 2012 [92] | 8 | 8 | 43.8 (6.58) | 44.8 (3.77) | 244.6 | √ |
| Nakamura 2013 [93] | 34 (14) | 51 (21) | 24.7 (5.5) | 23.9 (1.8) | 1.7 | √ |
| Neckelmann 2006 [94] | 12 (NA) | 12 (NA) | NA | NA | NA | √ |
| Nenadic 2012 [95] | 99 (42) | 113 (43) | 36.2 (11.2) | 32.4 (10.3) | NA | Χ |
| Nenadic 2015 [96] | 24 (12) | 49 (23) | 24.9 (3.1) | 23.8 (3) | 4.1 | √ |
| Oertel 2012 [97] | 31 (15) | 37 (20) | 38 (11.24) | 39.36 (9.97) | 164.5 | √ |
| Onay 2017 [98] | 20 (10) | 16 (9) | 36.5 (10.5) | 34.4 (9.1) | 128.4 | √ |
| Picado 2015 [99] | 20 (9) | 20 (8) | 35.9 (0.73) | 33.2 (6.6) | NA | √ |
| Rosa 2015 [100] | 32 (8) | 34 (15) | 28.2 (8.5) | 30.8 (8.3) | NA | √ |
| Salgado-Pineda 2004 [101] | 14 (7) | 14 (NA) | 25.1 (4.05) | 25.1 (3.32) | 17.5 | √ |
| Salgado-Pineda 2014 [102] | 14 (5) | 14 (5) | 37.29 (8.87) | 34.64 (5.96) | 168 | √ |
| Sapara 2016 [103] | 20 (4) | 20 (5) | 37.8 (7.85) | 35.3 (10.93) | NA | √ |

| | | | | | | |
|---|---|---|---|---|---|---|
| Schaufelberger 2007 [104] | 62 (18) | 94 (41) | 27.6 (8) | 30.2 (8.4) | 6.3 | √ |
| Segarra 2008 [105] | 28 (7) | 28 (7) | 27.3 (4.13) | 28.8 (5.36) | NA | √ |
| Singh 2014 [106] | 14 (3) | 14 (4) | 31.5 (9.4) | 27.2 (4.78) | 120.4 | √ |
| Singh 2018 [107] | 28 (16) | 28 (14) | 33.89 (9.34) | 31.44 (7.36) | 112.9 | √ |
| Spalthoff 2018 [108] | 51 (17) | 102 (33) | 35.18 (10.88) | 33.15 (9.6) | 105.6 | √ |
| Suazo 2014 [109] | 17 (7) | 13 (3) | 33.3 (10.48) | 30.9 (11.48) | 63.3 | x |
| Szendi 2017 [110] | 21 (10) | 13 (7) | 39 (NA) | 34 (NA) | 192 | √ |
| Tikasz 2019 [111] | 47 | 23 | 34.4 (9.6) | 31.9 (8.2) | NA | √ |
| Tomelleri 2009 [112] | 70 (25) | 79 (38) | 39.7 (10.94) | 40.3 (11.91) | 169.6 | √ |
| Torres 2016 [113] | 161 (50) | 151 (64) | 30.4 (8.3) | 30.6 (9) | 60 | √ |
| Tregellas 2007 [114] | 32 (11) | 32 (18) | 39.6 (8.8) | 35.3 (9.3) | NA | √ |
| van Tol 2014 [115] | 51 (7) | 51 (14) | 34 (11.4) | 36.1 (10.93) | 105.2 | √ |
| Venkatasubramanian 2010 [116] | 30 (9) | 27 (8) | 30.1 (8.3) | 27.4 (7) | 41.7 | X |
| Vijayakumari 2015 [117] | 41 (12) | 39 (7) | 27.3 (7.48) | 26.3 (5.85) | 24.5 | √ |
| Witthaus 2009 [118] | 23 (7) | 29 (12) | 26.4 (6.1) | 25.7 (5.2) | NA | √ |
| Yang 2019 [119] | 37 (16) | 28 (12) | 42 (8.44) | 40.5 (10.87) | 220.9 | √ |
| Yokoyama 2018 [120] | 60 (22) | 40 (11) | 38 (8.92) | 35.1 (7.6) | 152.3 | √ |
| Yue 2016 [121] | 20 (10) | 24 (11) | 24.5 (5.51) | 24.8 (6.11) | 22.9 | √ |
| Yuksel 2012 [122] | 43 (15) | 58 (20) | 38.7 (10.6) | 36.4 (10.5) | NA | √ |

| | | | | | | |
|---|---|---|---|---|---|---|
| Liu 2020 [123] | 75 (23) | 28 (9) | 28.35 (7.13) | 26.1 (5.57) | 80.4 | √ |
| Bose 2009 [124] | 34 | 33 | 40 (11) | 39.5 (12) | 144 | √ |
| Horn 2010 [125] | 20 (7) | 20 (7) | 30.1 (10) | 30.1 (NA) | NA | √ |
| McDonald 2005 [126] | 25 (7) | 52 (28) | 37.3 (10.2) | 39.3 (14.8) | 208.8 | √ |
| O'Daly 2007 [127] | 28 (3) | 32 (4) | 33 (10) | 34 (8) | 96 | √ |
| Palaniyappan 2012 [128] | 57 (7) | 41 (2) | 26.1 (7.49) | 28.04 (6.63) | NA | √ |
| Ediri 2020 [129] | 116 (67) | 116 (56) | 33.75 (9.5) | 33.41 (10.9) | 116.6 | √ |
| Adamu 2023 [130] | 72 (14) | 74 (23) | 38.17 (13.89) | 35.82 (11.58) | 192.36 | √ |
| Ma 2021 [131] | 52 (26) | 65 (32) | 26.67 (9.34) | 25.25 (4.07) | NA | √ |
| Burrer 2020 [132] | 60 (15) | 58 (24) | 30.5 (8.4) | 30.3 (8.2) | 100.8 | √ |
| Ruzic 2020 [133] | 106 (42) | 63 (33) | 30.3 (11.1) | 33 (4.45) | 132 | √ |
| Xie 2021 [134] | 30 (17) | 33 (20) | 30.3 (4.46) | 32.03 (7.31) | 21.36 | √ |
| Gou 2022 [135] | 70 (NA) | 43 (NA) | 29.93 (7.61) | 31.77 (7.17) | 63.79 | √ |
| Pinto 2023 [136] | 20 (7) | 20 (7) | 31.5 (10.3) | 31.5 (10.3) | 72 | √ |
| Lu 2024 [137] | 90 | 53 | 29.37 (7.13) | 31.17 (7.14) | 66.22 | X |
| Madeira 2020 [138] | 20 (7) | 20 (7) | 31.5 (10.3) | 31.5 (10.3) | 72 | √ |

Abbreviations: F, females, M, males, NA, not available, s.d standard deviation

**Supplementary Table 4. Demographic characteristics of patients with AUD**

| Study | Patients (F) | Healthy controls (F) | Mean age of patients (s.d) | Mean age of HCs (s.d) | Duration of use |
|---|---|---|---|---|---|
| vanHolst 2012 [139] | 36 (NA) | 54 (NA) | 43.2 (11) | 35.3 (10.1) | 140.28 |
| Mechtcheriakov 2007 [140] | 22 (8) | 22 (8) | 53.6 (NA) | 53.7 (NA) | NA |
| Wang 2016 [141] | 20 (NA) | 20 (NA) | 43.95 (6.3) | 40.5 (8.7) | NA |
| Chanraud 2007 [142] | 26 (NA) | 24(NA) | 47.7 (7.1) | 45 (6.72) | NA |
| Wiers 2015 [143] | 22 (NA) | 21 (NA) | 42.14 (6.2) | 41.95 (6.41) | 177.84 |
| Galandra 2018 [144] | 22 (9) | 18 (8) | 45.59 (7.99) | 44.83 (8.86) | 129.6 |
| Demirakca 2011 [145] | 50 (23) | 66 (32) | 46.6 (8.2) | 45 (10.1) | 148.8 |
| Chanraud 2009 [146] | 24 | 24 | 47.8 (7.7) | 45 (5.6) | NA |
| Nurmedov 2016 [147] | 24 (4) | 29 (6) | 40.79 (9.8) | 37.45 (10.87) | 92.4 |
| Segobin 2014 [148] | 19 (2) | 20 (NA) | 44.4 (6.07) | 46.7 (4.25) | 98.64 |
| Galandra 2021 [149] | 23 (NA) | 18 (NA) | 45.69 (7.82) | 44.83 (8.86) | 142.68 |
| Jang 2007 [150] | 20 | 20 | 43.5 (6) | 44.5 (7.4) | NA |
| Muller 2021 [151] | 23 (8) | 25 (7) | 42.1 (9.8) | 43.4 (10.2) | NA |

Abbreviations: F, females, M, males, NA, not available, s.d standard deviation

**Supplementary Table 5.** Disorder-specific meta-analytic results of MDD, SZ and AUD

| Contrast | Main brain regions | MNI coordinates | SDM Z |
|---|---|---|---|
| MDD > AUD | L precuneus | -2, -46, 38 | 6.469 |
| | R mid cingulate | 4, 24, 36 | 6.182 |
| | L insula | -44, -12, 4 | 6.353 |
| | L postcentral gyr | -38, -26, 52 | 4.733 |
| | R ant thalamus | 14, -34, 2 | 6.209 |
| | R sup temporal gyr | 54, -22, -6 | 5.274 |
| | R insula | 46, -24, 16 | 3.826 |
| | R gyr rectus | 4, 44, -20 | 4.372 |
| | R inf frontal gyr | 52, -58, 34 | 3.974 |
| | L thalamus | -12, -26, 6 | 5.041 |
| | R supramarginal gyr | 60, -38, 24 | 4.188 |
| | R anterior cingulate | -2, 24, -8 | 3.570 |
| SZ > AUD | L postcentral gyr | -46, -16, 48 | 5.925 |
| | R Precuneus | 2, -54, 28 | 4.704 |
| | L insula | -42, -12, 4 | 3.917 |
| | R lingual gyrus | 14, -34, 2 | 5.145 |
| | L SMA | -2, 0, 50 | 4.367 |
| | L anterior cingulate | 2, 40, 18 | 3.531 |
| MDD > SZ | L inf frontal gyr | -48, 20, 20 | 5.152 |
| | L thalamus | -2, -22, 8 | 5.386 |
| | R PHG | 24, -2, -30 | 4.255 |
| | R sup temporal gyr | 58, -38, 16 | 3.715 |
| | L sup temporal gyr | 2, 46, 42 | 4.170 |
| | R inf frontal gyr | 46, 4, 12 | 3.518 |
| | L pons | -12, -12, -20 | 3.249 |

| | | |
|---|---|---|
| L insula | -46, -12, 14 | 3.300 |
| R fusiform gyrus | 26, -72, -16 | -3.920 |

Abbreviations: AUD, Alcohol Use Disorder, ant, anterior, Cereb, cerebellum, Gyr, gyrus, HC, healthy controls, inf, inferior, L, left, MDD, major depressive disorder, MNI, Montreal Neurological Institute, PHG, parahippocampal gyrus, R, right, SMA, supplementary motor area, sup, superior, SZ, Schizophrenia.
